# A meta-analysis of cocaine use disorder treatment effectiveness

**DOI:** 10.1101/2020.06.09.20127142

**Authors:** Brandon S Bentzley, Summer S Han, Sherman C Stein, Sophie Neuner, Keith Humphreys, Kyle M Kampman, Casey H Halpern

## Abstract

**Background:** In both the U.S. and U.K., after a period of decline, prevalence of cocaine use has been increasing since 2012 and is now the second leading cause of overdose death from an illicit drug. However, psychosocial treatments for cocaine use disorders are limited, and no pharmacotherapy is approved by regulatory bodies in the U.S. or Europe. We performed a comprehensive meta-analysis to assess treatments’ impacts on cocaine use.

**Methods:** We performed a meta-analysis of clinical trials that included the word ‘cocaine’ in the title and were published between 31/12/1995 and 31/12/2017. All studies of outpatient adults with active cocaine use and reporting urinalysis results were included. Treatment approaches were clustered into 11 categories. Missing data were imputed using multiple imputation by chained equations. We calculated intention-to-treat log-odds ratios (OR) for the change in proportion of patients testing negative for cocaine at the end of each study and performed multivariate mixed-effects meta-regression. This study was prospectively registered on covidence.org on 31/12/2015, study 8731.

**Findings:** One hundred fifty-seven studies with 15,842 participants were included. Only contingency management was significantly associated with increased odds of testing negative for cocaine (OR of 2.13, 95% CI 1.62-2.80) and remained significant after all sensitivity analyses.

**Interpretation:** This meta-analysis is unique in its broad inclusivity of treatment types and trial designs over a two-decade period of investigation. Our results converge with focused meta-analyses on treatments for cocaine use disorders; thus, research efforts and policy changes that expanded implementation of contingency management programs are expected to reduce cocaine use in active users and the associated individual, community, and societal burdens associated.

**Funding:** None

**Panel:** *Evidence before this study:* Before undertaking this study, we examined all cocaine use disorder treatment reviews in Cochrane Collaboration as well as all meta-analyses indexed on PubMed (search term = “cocaine” and article type = Meta-analysis). We identified meta-analyses of several treatments for cocaine use disorders that were negative or indeterminant, including anticonvulsants, antidepressants, antipsychotics, acupuncture, disulfiram, dopamine agonists, opioids, and psychostimulants. Meta-analyses of psychosocial interventions showed variable effect sizes with large heterogeneity between approaches. Meta-analyses of contingency management indicated efficacy in reducing cocaine use, but these have been limited to specific subpopulations or to controlled studies. We expanded our scope beyond prior investigations to comprehensively assess all treatment categories simultaneously across all study types with the aim of increasing our sensitivity for detecting an effective treatment for cocaine use disorders in an otherwise largely negative evidence base.

*Added value of this study:* Our findings indicate robust effectiveness of contingency management approaches in reducing cocaine use. Other treatment categories were either negative or failed sensitivity testing. This finding highlights the inaccuracy of the common notion that there is no effective treatment for cocaine use disorder.

*Implications of all the available evidence:* Based on our study, contingency management is an effective treatment for cocaine use disorder. Cocaine use and its associated adverse effects could be significantly reduced in patients suffering from cocaine use disorder through expanded implementation of contingency management programs.

## Introduction

After years of decline, the prevalence of cocaine use has been increasing since 2012, and now, following opioids, cocaine is the second leading cause of overdose death from an illicit drug in the United States (US)^1,2^ and in the United Kingdom^3^. It takes a particular toll on certain vulnerable populations, for example being the leading cause of overdose deaths among Black persons^4^. However, treatments for cocaine use disorders are limited, and, despite hundreds of clinical trials conducted over several decades, there continues to be no pharmacotherapy approved by government agencies in the U.S. or Europe. This is in contrast to opioid agonist therapy for treatment of opioid use disorder^5–7^, naltrexone for alcohol use disorder^8^, or varenicline to treat tobacco use disorders^9,10^.

Absence of an efficacious standard treatment for cocaine use disorders hampers clinical treatment, and with no guiding prototype available, development of new treatments has proven challenging. Further, current understanding of the pathophysiology of cocaine use disorders remains insufficient for developing efficacious pharmacological treatments. Numerous meta-analyses have attempted to search for an efficacy signal by pooling results from multiple clinical trials. Unfortunately, meta-analytic investigations have shown no improvement of outcomes for anticonvulsants^11–13^, antidepressants^14^, antipsychotics^15–17^, acupuncture^18^, disulfiram^19^, dopamine agonists^20^, opioids^21^, and psychostimulants^22–24^. Meta-analyses of psychosocial interventions have shown variable effect sizes given the heterogeneity of approaches^21,25,26^. Meta-analyses of contingency management – positive reinforcement of drug abstinence – have indicated efficacy; however, these studies have been limited to unique populations, to those with specific comparison groups^21,27–29^ or to exclusively studies of contingency management^30^. Further, leaders in the field continue to classify contingency management as a treatment with limited effectiveness^31^, making its comparative role in treating cocaine use disorder unclear. Herein, we performed a comprehensive meta-analysis of all treatments for cocaine use disorders published over 22 years, inclusive of all clinical trials and cocaine-using populations, to determine which treatment approaches, if any, are associated with a reduction in cocaine use.

## Methods

### Search strategy and selection criteria

This meta-analysis was prospectively registered on covidence.org on December 31, 2015, study 8731. Covidence.org was used to store search results, identify duplicates, and track screening decisions. The PubMed database was searched for clinical trials with the word “cocaine” in the title that were published between December 31, 1995 and December 31, 2015. This search was temporally expanded and repeated on December 31, 2016 and December 31, 2017 to update the analysis with relevant studies. Only English language articles were included (this excluded 6 of 772 abstracts, with only 2 of these 6 describing small studies relevant to this meta-analysis). The exact search string was as follows: cocaine[title] AND (Clinical Trial[ptyp] AND “loattrfull text"[sb] AND (“1995/12/31"[PDAT]: “2017/12/31"[PDAT]) AND English[lang]). Additionally, all references within Cochrane meta-analyses on treatments for cocaine use disorders and additional references identified during full text screening were included in abstract screening.

All clinical trial designs were included as long as the primary goal of the study was to test efficacy of a treatment for reducing cocaine use. Participants had to be 18 years of age or older with active cocaine use at baseline by self-report or urinalysis testing. Studies were excluded if >25% participants were not active cocaine users by self-report, or >80% of participants tested negative for cocaine at baseline. Only studies that reported treatment group size, treatment duration, retention rates, and treatment outcomes as urinalysis testing for cocaine metabolites were included. Studies that reported treatment outcomes only as urinalysis results pooled across multiple drugs were excluded, i.e. urinalysis results not reporting the specific proportion testing negative/positive for cocaine metabolites.

Study authors were not contacted, nor were unpublished data sought. Data included in the meta-analysis was extracted at the summary estimate level. BB performed the search, full text screening, data extraction, and data analysis. SN and BB screened all abstracts and references. SH and BB performed data analysis. Conflicts regarding inclusion were resolved by re-consideration by BB; however, no conflicts arose. A detailed outline of the study protocol can be found online (webappendix p 2-5).

### Outcome measures

The start and end of treatment was defined as the first and last timepoint participants were respectively exposed to the treatment; post-treatment data were not included. The primary outcome was defined as the intention-to-treat (ITT) log-odds ratio (OR) of testing negative for cocaine by urinalysis at the end of the treatment period compared to baseline. Baseline urinalysis data were either reported directly, inferred based on a requirement of testing positive for cocaine for study entry, or estimated based on the urinalysis results during the first week of treatment. The type of baseline data reported was coded as a dummy variable and included in the statistical analysis. For studies reporting multiple baseline types, direct baseline testing was preferred over testing positive at screening, which was preferred over estimation based on the first week of treatment. Outcome urinalysis data were reported either directly as ITT or calculated based on retention rate and non-ITT outcome. End of treatment urinalysis was either reported at the last treatment timepoint or as mean urinalysis for the entire treatment period. The type of outcome data reported was coded as a dummy variable and included in the statistical analysis. For studies reporting multiple outcome types, data from the last treatment timepoint were preferred over mean urinalysis results across the treatment period, and direct ITT reporting was preferred over calculating ITT outcomes based on retention and non-ITT outcomes.

### Data extraction

All search results were imported from PubMed XLM output into Covidence.org with duplicates automatically being removed during importation. Two reviewers (BB and SN) independently reviewed references and abstracts. If both reviewers agreed that the trial did or did not meet eligibility criteria, it was included or excluded respectively. We obtained the full text of all remaining articles and used the same eligibility criteria to determine which, if any, to exclude at this stage. Any disagreements were solved via discussion and were ultimately under the discretion of BB and CH. BB read each full text article, determined if it met inclusion criteria, and extracted data to a Microsoft Excel database backed up continuously to offsite storage. Data extraction was completed in two iterations with the second iteration ensuring fidelity of the first. In addition to urinalysis outcome and retention data, information extracted included study characteristics (lead author, publication year, double-blindedness, randomization, multi-sitedness), participant characteristics (age, gender, years of cocaine use, self-reported days of cocaine use per week, Addiction Severity Index drug composite scale), and intervention details (treatment categories, specific treatment, dose, duration of treatment). All treatments that participants were simultaneously exposed to in a treatment arm within a study were coded by category: Psychotherapy, Contingency Management, Placebo, Opioid, Psychostimulants, Anticonvulsants, Dopamine Agonists, Antidepressants, Antipsychotics, Miscellaneous Medications (medication that did not fit in other medication categories), or Other (non-medication that does not fit in any treatment category). For example, a single treatment group may have been treated with fluoxetine, cognitive behavioral therapy, and methadone concomitantly, and this treatment group would be coded with the following treatment categories: Antidepressants, Psychotherapy, and Opioids.

### Statistical analysis

All statistical analyses were performed with R^32^ 3.3.2 using the Metafor^33^ (meta-regression) and MICE^34^ (data imputation) packages. All R script with associated text output can be found in the webappendix (pp 6-56). The escalc function within the Metafor package was used to calculate log odds ratio (*y*_*i*_=*ln*[(*p*/1-*p*)/(*q*/1-*q*)]) and variability (*v*_*i*_) for each treatment group based on the group size and proportion of urinalysis results that were negative for cocaine metabolites at the start (*q*) and end of treatment (*p*). This method takes into account the number of participants in each treatment group with decreasing variability with increasing group size.

The multiple imputation by chained equations (MICE) package in R was used to impute missing data. MICE is a robust method of data imputation that creates multiple predictions for each missing value and then pools the results to account for uncertainty in the imputations and estimate accurate standard errors^35^. Predictive mean matching was used to impute missing baseline urinalysis data, duration of treatment, age, proportion male, years of cocaine use, and addiction severity index (ASI)^36^ drug subscale. Briefly, predictive mean matching uses linear regression with all observed data from all variables to build a predictor matrix. The algorithm then finds cases with observed data in which the predicted value of the observed data point is proximal to the predicted value of the missing data point. It selects 5 donors from the closest matches, randomly samples an observed value from one of the donors and uses this to impute the missing data point. As imputed data is always an observed value within the data set, it always meets boundary criteria and has the same distribution. The type of baseline urinalysis data (inferred by positive cocaine urinalysis being required at study entry, measured at baseline, or measured during the first week) was imputed using a Bayesian polytomous regression model built from all observed data. Data imputation was run 5 times to build 5 separate imputed data sets.

In our primary analyses, we constructed a multilevel random-effects model with treatment groups nested within studies and studies nested within the first author, e.g. multiple studies conducted by Jill Smith would be nested together. The interclass correlation coefficient was used to assess effect of nesting and Higgins I^2^ was used to estimate heterogeneity. Collinearity was defined by a variance inflation factor (VIF) > 5. Statistical significance was set at 0.05. Multilevel mixed-effects meta-regression was conducted for the dataset composed of studies with complete data for baseline urinalysis results, treatment duration, proportion male, and mean age. ASI was not included in the non-imputed analyses, as most studies did no report it. This analysis was then repeated for the 5 imputed data sets, and results of these 5 analyses were pooled. These 2 analyses (multi-level without imputed data and pooled results of 5 multi-level models with imputed data) served as the primary analyses. Sensitivity analyses were conducted by examining the results of several other random effects models: with treatment factors only, with retention rate as a covariate, single level modelling, and assessing the results of each of the 5 imputed data sets on an individual basis.

## Results

In total, we included 113 studies with 402 treatment groups and 15,842 participants in the meta-analysis. See figure 1 for the PRISMA^37^ diagram, which includes information on the number of studies assessed for eligibility, the number ineligible, and a list reasons for exclusion.

**Figure 1:**
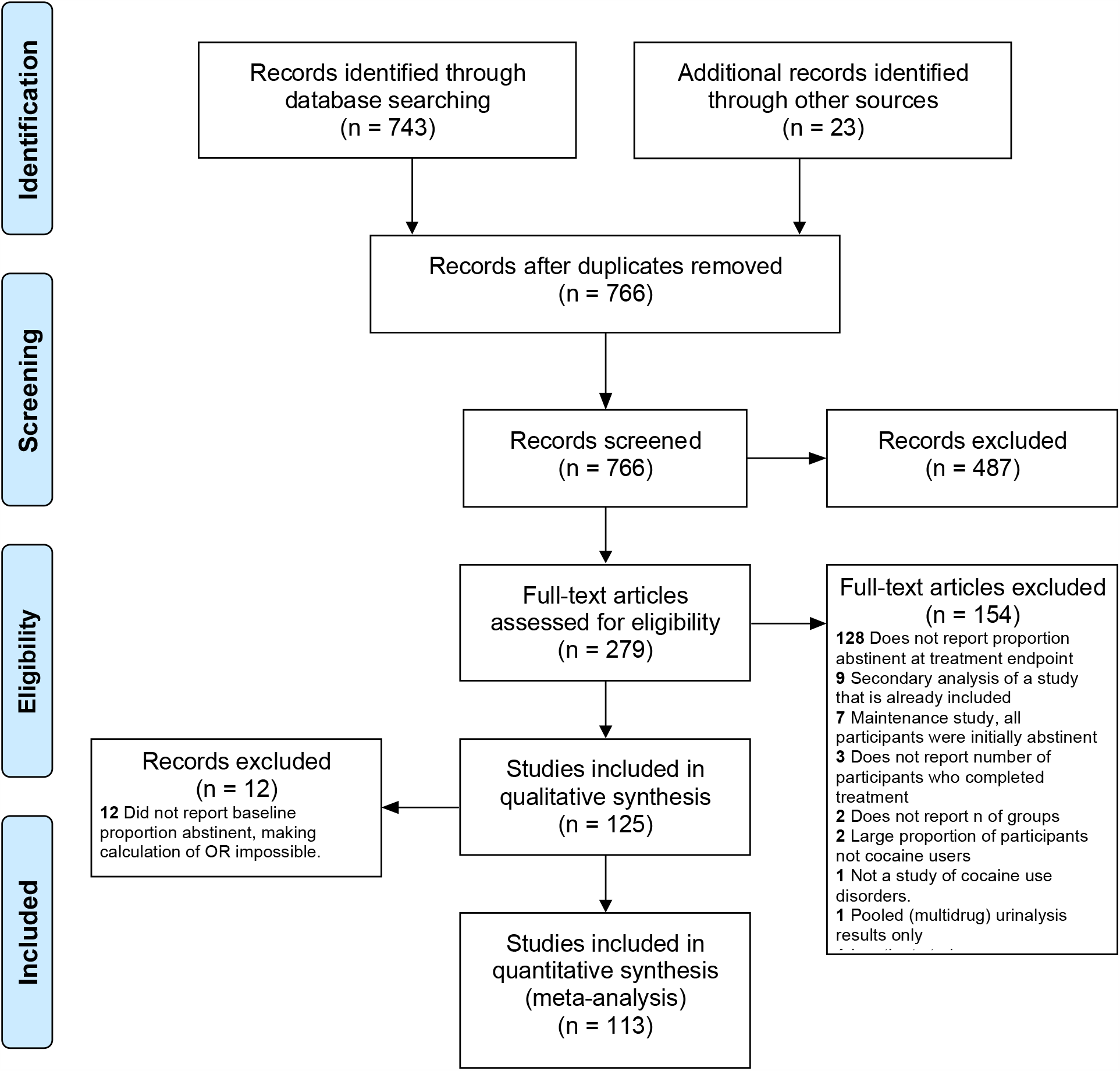
Study selection.

Forest plots with unadjusted outcomes for all studies with baseline data are included by treatment category in the webappendix pp 57-76. Table 1 includes summary statistics for each treatment category and covariates. Table 2 includes main outcome measures for the primary analyses: multi-level model with covariates with and without imputed data. For the primary multi-level analysis with covariates, several treatments were significantly associated with a significant increase in the ITT odds of producing a negative urinalysis in the non-imputed dataset; however, only contingency management was significant for both imputed (OR = 2.13, 95% CI = 1.62-2.80) and non-imputed (OR = 2.09, 95% CI = 1.59-2.75) datasets. This significance persisted across all sensitivity testing (Table 3). Notably, placebo was not associated with a significant change in the ITT log odds of producing a negative urinalysis for the imputed (OR = 1.03, 95% CI = 0.59-1.80) nor non-imputed (OR = 1.48, 95% CI = 0.86-2.53) dataset, including all sensitivity testing (Table 3). When completion rate was added as a covariate to the primary non-imputed multi-level model, only contingency management remained significantly associated with outcomes (OR = 2.06, 95% CI = 1.53-2.77).

**Table 1:** Summary statistics for each treatment category and covariates.

|  | Treatment groups |  |  |  |  |  |  |  |  |  |  |
| --- | --- | --- | --- | --- | --- | --- | --- | --- | --- | --- | --- |
| Characteristic | Anticonvulsants | Antidepressants | Antipsychotics | Contingency Management | Dopamine Agonists | Misc Meds | Opioids | Other | Placebo | Psychostimulants | Psychotherapy |
| N Treatment Groups | 20 | 18 | 12 | 88 | 20 | 57 | 159 | 27 | 96 | 24 | 331 |
| N Participants | 683 | 574 | 281 | 3301 | 710 | 1796 | 5139 | 1873 | 3381 | 702 | 13248 |
| Mean N Participants (STD) | 34.15 (4.62) | 31.89 (3.73) | 23.42 (3.17) | 37.51 (2.72) | 35.50 (4.29) | 31.51 (3.75) | 32.32 (1.48) | 69.37 (15.80) | 35.22 (2.59) | 29.25 (3.47) | 40.02 (2.02) |
| Mean Age (STD, #treatment groups missing) | 37.21 (5.47,0) | 36.35 (2.33,0) | 39.77 (4.78,0) | 36.78 (4.57,1) | 38.34 (4.67,0) | 39.93 (4.68,1) | 36.44 (3.86,0) | 37.77 (3.79,3) | 36.05 (5.21,1) | 37.39 (3.07,0) | 36.52 (4.26,4) |
| Proportion Male | 0.75 (0.15,0) | 0.70 (0.26,1) | 0.75 (0.21,0) | 0.55 (0.21,2) | 0.80 (0.11,0) | 0.71 (0.13,2) | 0.67 (0.10,4) | 0.69 (0.16,0) | 0.79 (0.17,1) | 0.74 (0.12,1) | 0.79 (0.18,7) |
| Mean days per week cocaine use | 3.97 (0.95,3) | 3.08 (1.04,1) | 2.58 (0.84,7) | 3.85 (0.68,33) | 3.25 (0.60,6) | 4.00 (1.10,7) | 2.82 (0.89,43) | 3.47 (0.44,7) | 3.36 (0.87,18) | 2.90 (0.44,9) | 3.35 (0.97,110) |
| Mean years cocaine use | 10.82 (2.44,11) | 9.55 (1.65,14) | 14.25 (4.46,9) | 8.00 (1.69,37) | 9.80 (3.06,3) | 11.79 (3.38,15) | 8.81 (3.71,74) | 11.18 (4.56,11) | 10.55 (3.76,39) | 12.28 (2.66,11) | 11.00 (4.86,128) |
| Mean ASI Drug | 0.30 (0.07,12) | 0.33 (0.08,15) | 0.24 (0.01,9) | 0.27 (0.05,56) | 0.23 (0.03,12) | 0.24 (0.03,44) | 0.29 (0.00,121) | 0.30 (0.05,18) | 0.22 (0.01,69) | 0.24 (0.01,20) | 0.26 (0.18,216) |
| Proportion cocaine free at start | 0.17 (0.20,3) | 0.07 (0.15,3) | 0.31 (0.24,1) | 0.14 (0.15,19) | 0.24 (0.18,2) | 0.06 (0.09,16) | 0.17 (0.15,17) | 0.09 (0.14,6) | 0.11 (0.17,18) | 0.25 (0.16,5) | 0.13 (0.14,71) |
| Mean weeks treatment duration | 11.07 (1.59,0) | 12.40 (1.53,0) | 13.27 (6.82,0) | 16.15 (7.05,0) | 11.07 (4.80,0) | 10.05 (2.23,0) | 14.86 (6.64,0) | 14.77 (6.37,0) | 10.33 (2.87,0) | 11.56 (4.42,0) | 12.96 (7.01,0) |
| Proportion completed treatment | 0.59 (0.21,0) | 0.45 (0.20,0) | 0.49 (0.24,0) | 0.73 (0.15,5) | 0.41 (0.13,0) | 0.57 (0.25,0) | 0.63 (0.17,2) | 0.72 (0.19,0) | 0.69 (0.18,0) | 0.50 (0.20,0) | 0.54 (0.27,7) |
| Proportion completed treatment and cocaine free | 0.25 (0.15,0) | 0.24 (0.12,0) | 0.20 (0.17,0) | 0.40 (0.19,0) | 0.20 (0.15,0) | 0.23 (0.17,0) | 0.24 (0.11,0) | 0.28 (0.17,0) | 0.29 (0.17,0) | 0.21 (0.10,0) | 0.30 (0.19,0) |

**Table 2:** Outcomes for primary analyses.

|  | Without imputed data |  |  |  |  | With imputed data |  |  |  |  |
| --- | --- | --- | --- | --- | --- | --- | --- | --- | --- | --- |
| Treatment Category | N studies | N treatment groups | N participants | OR | 95% CI | N studies | N treatment groups | N participants | OR | 95% CI |
| Anticonvulsants | 14 | 17 | 570 | 2.32 | 1.15-4.67 | 16 | 20 | 683 | 1.39 | 0.59-3.29 |
| Antidepressants | 8 | 15 | 482 | 1.96 | 0.91-4.25 | 11 | 18 | 574 | 1.36 | 0.63-2.91 |
| Antipsychotics | 8 | 11 | 241 | 1.59 | 0.68-3.71 | 9 | 12 | 281 | 1.02 | 0.41-2.53 |
| Contingency Management | 38 | 69 | 2744 | 2.09 | 1.59-2.75 | 50 | 88 | 3301 | 2.13 | 1.62-2.80 |
| Dopamine Agonists | 10 | 18 | 645 | 1.55 | 0.80-3.00 | 12 | 20 | 710 | 1.04 | 0.51-2.14 |
| Misc Meds | 26 | 41 | 1500 | 1.64 | 0.88-3.06 | 37 | 57 | 1796 | 1.14 | 0.60-2.17 |
| Opioids | 49 | 142 | 4686 | 2.32 | 1.54-3.49 | 55 | 159 | 5139 | 1.70 | 0.98-2.94 |
| Other | 12 | 21 | 1577 | 2.19 | 1.07-4.49 | 15 | 27 | 1873 | 1.35 | 0.72-2.53 |
| Placebo | 65 | 78 | 2889 | 1.48 | 0.86-2.53 | 80 | 96 | 3381 | 1.03 | 0.59-1.80 |
| Psychostimulants | 13 | 19 | 645 | 2.48 | 1.27-4.85 | 17 | 24 | 702 | 1.74 | 0.81-3.74 |
| Psychotherapy | 98 | 260 | 10808 | 1.08 | 0.74-1.58 | 131 | 331 | 13248 | 1.18 | 0.81-1.73 |

**Table 3:** Sensitivity analyses.

|  | Analysis |  |  |  |  |  |  |  |  |  |  |  |  |  |  |  |  |
| --- | --- | --- | --- | --- | --- | --- | --- | --- | --- | --- | --- | --- | --- | --- | --- | --- | --- |
| Variable | 1 | 2 | 3 | 4* | 5 | 6 | 7 | 8 | 9 | 10 | 11* | 12 | 13 | 14 | 15 | 16 | 17 |
| Anticonvulsants |  | + |  | + |  |  |  |  |  |  |  |  |  |  |  |  |  |
| Antidepressants | +++ |  |  |  |  |  |  |  |  |  |  |  |  |  |  |  |  |
| Antipsychotics |  |  |  |  |  |  |  |  |  |  |  |  |  |  |  |  |  |
| Contingency Management | ++ | +++ | +++ | +++ | +++ | +++ | +++ | +++ | +++ | +++ | +++ | +++ | +++ | +++ | +++ | +++ | +++ |
| Dopamine Agonists |  |  |  |  |  |  |  |  |  |  |  |  |  |  |  |  |  |
| Misc Meds | + |  |  |  |  |  |  |  |  |  |  |  |  |  |  |  |  |
| Opioids | +++ | +++ | + | +++ | + | + | + | ++ |  | ++ |  |  |  |  |  |  |  |
| Other | + | ++ | ++ | + | ++ |  |  |  |  |  |  |  |  |  |  |  |  |
| Placebo |  |  |  |  |  |  |  |  |  |  |  |  |  |  |  |  |  |
| Psychostimulants |  |  | + | ++ | + | + |  |  |  |  |  |  |  |  |  |  |  |
| Psychotherapy |  |  |  |  |  |  |  |  |  |  |  |  |  |  |  |  |  |
| Double Blind | NA | + |  | + |  |  |  |  |  |  |  |  |  |  |  |  |  |
| Multisite | NA |  |  |  |  |  |  |  |  |  |  |  |  |  |  |  |  |
| Randomized | NA |  |  |  |  |  |  |  |  |  |  |  |  |  |  |  |  |
| Baseline Type 2 | NA | +++ | +++ | +++ | +++ | +++ | +++ | +++ | +++ | +++ | ++ | +++ | +++ | +++ | +++ | +++ | +++ |
| Baseline Type 3 | NA | +++ | +++ | +++ | +++ |  | +++ | +++ | +++ |  |  |  | +++ | ++ | +++ |  |  |
| Outcome Type 2 | NA | + |  |  |  |  | + | + |  |  |  |  |  |  |  |  |  |
| Outcome Type 3 | NA | ++ | ++ | + | ++ |  | +++ | +++ |  | ++ |  |  | + |  |  | ++ |  |
| Outcome Type 4 | NA | +++ |  |  |  |  | +++ | +++ | + | + |  | + | +++ | +++ | +++ | ++ |  |
| Duration | NA | + | + |  | + | + |  |  | + |  |  | ++ |  |  | ++ | + |  |
| %Male | NA | ++ | + |  | + | + |  | ++ |  |  |  | + |  | + |  |  |  |
| Age | NA |  |  |  |  |  |  |  |  |  |  |  |  |  |  |  |  |
| Baseline days per week cocaine use | NA |  |  |  |  |  |  | + |  |  |  |  |  | + | ++ |  |  |
| Year published | NA | ++ | + |  | + |  |  |  |  |  |  |  |  |  |  |  |  |
| ASI Drug | NA | NA | NA | NA | NA |  |  |  |  |  |  |  |  |  |  |  |  |
| Completion rate | NA | NA | +++ | NA | +++ | NA | NA | NA | NA | NA | NA | +++ | +++ | +++ | +++ | +++ | +++ |

1 = Treatments only, 2 = Treatments with covariates, 3 = Treatments with covariates + completion rate, 4 = Multilevel: Treatments with covariates, 5 = Multilevel: Treatments with covariates + completion rate, 6-10 = Multilevel: Treatments with covariates imputed-Models 1-5, 11 = Multilevel: Treatments with covariates imputed-pooled, 12-16 = Multilevel: Treatments with covariates imputed + completion rate-Models 1-5, 17 = Multilevel: Treatments with covariates imputed + completion rate-pooled, \*primary analysis (4 and 11), +p<0.05, ++p<0.01, +++p<0.002, NA not applicable, Baseline Types are all in comparison to Baseline Type 1 = all participants had to test positive for cocaine at screening for study inclusion, Baseline Type 2 = proportion of participants urinalysis positive for cocaine reported at baseline, Baseline Type 3 = proportion of participants urinalysis positive for cocaine reported during first week of treatment, Outcome Types are all in comparison to Outcome Type 1 = proportion of participants reported urinalysis positive reported as ITT during last week of treatment, Outcome Type 2 = ITT proportion of participants reported urinalysis positive calculated from retention rate and outcome during last week of treatment, Outcome Type 3 = mean proportion urinalyses positive reported as ITT during treatment, Outcome Type 4 = ITT mean proportion urinalyses positive calculated from retention rate and outcome during treatment.

Baseline data type was the only covariate significantly associated with outcome for the primary multi-level analysis with covariates, with baseline urinalysis results reported prior to treatment being associated with lower ITT odds of producing a negative urinalysis for both imputed (OR = 0.26, 95% CI = 0.13-0.55) and non-imputed (OR = 0.13, 95% CI = 0.08-0.20) datasets when compared to the reference of testing positive for cocaine by urinalysis as a requirement for study entry. This is in contrast to baseline data type being reported as the urinalysis results for week 1 of treatment, which was significant for the non-imputed (OR = 0.11, 95% CI = 0.06-0.20) but not for the imputed (OR = 0.42, 95% CI = 0.03-5.45) dataset.

Heterogeneity (I^2^) was 80.96% in the base random effects model without any treatment category factors nor covariates, 41.79% with all treatment categories and covariates except completion rate, 22.57% when completion rate was also included as a covariate, 58.54% in the multi-level model with all treatment categories and covariates except completion rate, and 49.40% when completion rate was also included as a covariate. I^2^ ranged from 72.69% to 76.72% across the five imputed data sets without completion rate and 68.35% to 72.78% for the five studies with completion rate as a covariate. The interclass correlational coefficient was 0.498 for the multilevel model without completion rate and 0.392 with completion rate as a covariate. The interclass correlational coefficient ranged from 0.222 to 0.608 without completion rate and 0.107 to 0.548 with completion rate as a covariate. Examination of funnel plots across models and across treatment categories indicated low risk of bias (webappendix pp 77-79). Collinearity was not detected for any treatment category or covariate by VIF.

## Discussion

This meta-analysis was constructed to maximize inclusivity to detect a broad treatment category that is both generalizable and effective in reducing cocaine use. Although several categories demonstrated efficacy in the non-imputed data set, when data were imputed to include the complete data set, only contingency management was consistently associated with a significant reduction in urinalysis-confirmed cocaine use. Other treatment categories were not significantly associated with this outcome. This is in contrast to placebo, which was consistently not associated with a significant change in objective cocaine use in any of the primary or sensitivity analyses.

Our *a priori* hypothesis, based on results of the majority of previously published meta-analyses, was that no treatment category would have a significant association with objective cocaine use. However, the positive association between contingency management treatment approaches and a significant reduction in objective cocaine use is not entirely surprising. Prior meta-analyses of contingency management for reducing cocaine use have suggested efficacy in particular clinical populations^21,27,28^, and a recent high-quality meta-analysis of psychosocial treatments with comparison groups for treatment of cocaine use disorders found contingency management the most effective treatment^29^. Moreover, large-scale implementation of contingency management for treatment of substance use disorders in the Department of Veterans Affairs (VA) has demonstrated both clinical effectiveness, similar to what has been reported in controlled trials, and low cost^38^. Given that the VA is the largest integrated provider of addiction services in the US^39^ and the results of our study reported herein, serious consideration for implementation of contingency management on a national level or within other major healthcare systems in the US is warranted.

After contingency management, opioids were the treatment category that demonstrated efficacy in the most analyses. Opioid agonist therapies were significantly associated with a reduction in cocaine use in 9 of 11 analyses that did not include completion rate as a covariate and 2 of 8 analyses that included completion rate, indicating that the efficacy of opioids in reducing ITT cocaine use was enhanced through an increase in retention rate. Notably, all studies that included opioids (buprenorphine and methadone) as a treatment were in populations with concomitant opioid use disorders. These medications have a known significant effect on increasing retention rate^7^ and similarly to our report here, the only other meta-analysis on the treatment of cocaine use using opioids did not find an a significant effect outside of this enhanced retention^21^.

Our analysis did not reveal a significant association between psychotherapy and cocaine use. Meta-analyses of psychosocial interventions have shown variable effect sizes that have been attributed to the heterogeneity of approaches^21,25,26^. Similarly, our analysis did not take into account the type or dose (session length and frequency) of psychotherapy provided. Over three-fourths (331 of 402) treatment groups incorporated some form of psychotherapy, and open-label as well as non-controlled study designs were included. Hence, if a general effect of psychotherapy on cocaine use were present, it would have likely been detected. However, our approach cannot rule out efficacy of specific approaches or doses. This is representative of the major strength – broadly inclusive with increased likelihood of detecting an effect that is consistently present in a treatment category – and weakness – inability to resolve effect size of specific treatments nor identify potential efficacy of a specific treatment within a broad treatment category.

In addition to our approach being biased to detect a response that is consistently present within a treatment category, as opposed to only within a specific treatment within a category, there are at least two other unique aspects of our analyses – the inclusion of all eligible studies regardless of quality and effect sizes calculated from baseline to treatment end instead of measuring treatment efficacy in comparison to a control group, even when one was present. Both of these approaches were done to maximize the sensitivity of detecting an effective treatment; however, at the expense of increasing the probability of a type 1 error. We compensate for this by reporting the results of all sensitivity analyses and limiting our conclusion to the significant effect of contingency management on cocaine use, the only treatment category positive for both primary analyses and the only treatment category positive for all sensitivity analyses. Furthermore, study quality was not found to be related to outcomes, so inclusion of low-quality studies is unlikely to have biased results.

In conclusion, our comprehensive analyses here indicate that contingency management approaches significantly reduce cocaine use. Thus, there is no case for therapeutic nihilism regarding cocaine use disorder. Prioritizing implementation research that informs healthcare systems in viable adoption approaches and assesses associated effectiveness would likely produce greater public health benefit than additional efforts to determine efficacy of contingency management approaches.

## Data Availability

The article is a meta-analysis of published works.

## Author Contributions

Brandon S Bentzley: study design, literature search, data collection, data analysis, data interpretation, figures, writing

Summer S Han: study design, data analysis, data interpretation, writing Sherman Stein: study design, data collection, data analysis, data interpretation Sophie Neuner: literature search, data collection,

Keith Humphreys: data interpretation, writing Kyle M Kampman: data interpretation, writing

Casey H Halpern: study design, data analysis, data interpretation, writing

## Ethics committee approval

This was a meta-analysis and required no involvement of an ethics committee.

